# Faster post-malnutrition weight gain during childhood is associated with risk of non-communicable disease in adult survivors of severe malnutrition

**DOI:** 10.1101/2022.03.02.22271635

**Authors:** Debbie S. Thompson, Kimberley McKenzie, Charles Opondo, Michael S. Boyne, Natasha Lelijveld, Jonathan Wells, Tim J. Cole, Kenneth Anujuo, Mubarek Abera, Melkamu Berhane, Marko Kerac, CHANGE Study Collaborators Group, Asha Badaloo

## Abstract

**Background:** Nutritional rehabilitation during severe malnutrition (SM) aims to rapidly restore body size and minimize poor short-term outcomes. We hypothesized that too rapid weight gain during and after treatment might however predispose to cardiometabolic risk in adult life.

**Methods:** Weight and height during hospitalization and one year post-hospitalization were abstracted from hospital records of children who survived SM. Six definitions of post-malnutrition weight gain/growth were analysed as continuous variables, quintiles and latent classes in age-sex and minimum weight-for-age z-scores-adjusted regression models against adult anthropometry, body composition (DEXA), blood pressure, blood glucose, insulin, and lipids.

**Results:** 60% of 278 participants were male, mean (SD) age 28.2 (7.7) years, mean (SD) BMI 23.6 (5.2) kg/m^2^. Mean admission age for SM was 10.9 months (range 0.3-36.3 months) and 207/270 (77%) were wasted (weight-for-height z-score<-2). During childhood, mean rehabilitation weight gain (SD) was 10.1(3.8) g/kg/day and 0.8(0.5) g/kg/day in the first year post-hospitalization. Rehabilitation weight gain >12.9 g/kg/day was associated with higher adult BMI (difference=0.5kg/m^2^, 95%CI: 0.1-0.9, *p* = 0.02), waist circumference (difference=1.4cm, 95%CI: 0.4-2.4, *p*=0.005), fat mass (difference = 1.1kg, 95%CI: 0.2-2, *p*=0.02), fat mass index (difference=0.32, 95%CI: -0.0001-0, *p*=0.05), and android fat mass (difference=0.09 kg, 95%CI: 0.01-0.2, *p*=0.03). Rehabilitation (g/month) and post-hospitalization (g/kg/month) weight gain were associated with greater lean mass (difference = 0.7 kg, 95% CI: 0.1, 1.3, *p* = 0.02) (difference=1.3kg, 95% CI: 0.3-2.4, *p*=0.015) respectively.

**Conclusion:** Rehabilitation weight gain exceeding 13g/kg/day was associated with adult adiposity in young, normal-weight adult SM survivors. This raises questions around existing malnutrition weight gain targets and warrants further studies exploring optimal post-malnutrition growth.

## Introduction

Severe malnutrition (SM) remains an important cause of mortality globally, and malnutrition contributes to the deaths of 500,000 children under the age of 5 years annually [1]. As global efforts improve the management of these children, alongside increased availability of ready to use therapeutic foods, the mortality from severe malnutrition is declining in some settings. However, while the short-term outcomes of severe childhood malnutrition may be improving, the long-term consequences are less clear. Although there is evidence of an association between prenatal undernutrition and later risk of chronic non-communicable diseases (NCDs) [2, 3], much of this evidence is from developed countries. Additionally, evidence of similar links between early postnatal malnutrition and later NCDs is recent and limited [4-6], and potential mechanisms are yet to be identified.

Most malnutrition treatment programs treat the attainment of normal weight-for-height as an indicator of recovery and aim for this to occur as rapidly as possible. To this end, energy dense feeds are usually provided during the rehabilitation stage of treatment of severe malnutrition; children are fed increasing amounts of energy and protein-enriched feeds to achieve a normal body weight. The recommended energy intake during the three day transition period is 100-135 kcal/kg/day [7] followed by intake of 150-225 kcal/kg/d during the rehabilitation (“catch-up growth”) period, the goal being a weight gain of ≥ 10 g/kg/day as recommended by the World Health Organization (WHO) [8]. However, it is unclear whether better short-term outcomes are occurring at the expense of longer-term outcomes. The thrifty phenotype hypothesis proposes that poor infant growth is a risk factor for the subsequent development of type 2 diabetes and the metabolic syndrome due to the effects of poor early-life nutrition [9]. In such “metabolically thrifty” individuals, faster weight gain in the plastic developmental period of infancy could pose an additional risk for later NCDs [10].

Many countries with high levels of early childhood malnutrition also have high burdens of obesity and NCDs including hypertension, type 2 diabetes and cardiovascular disease [11]. Data from the WHO indicate that NCDs are the leading causes of death and disability globally, killing more than three in five people worldwide and responsible for more than half the global burden of disease [12]. NCDs also cause and perpetuate poverty while hindering economic development in low- and middle-income countries (LMICs) as they struggle with the economic burden of providing treatment for individuals with NCDs, thus recapitulating the cycle.

This study aims to explore rehabilitation and post-hospitalization weight gain and growth in children who were hospitalized with severe malnutrition, and to investigate the associations between weight gain during and after rehabilitation and adult obesity, body composition and NCD risk. We hypothesize that faster rates of growth during nutritional rehabilitation are associated with later NCD risk.

## Methods

Setting: We conducted secondary analysis of data collected from members of the LION (Long-term Implications of Nutrition) Cohort. The LION Cohort was retrospectively assembled and from individuals who were admitted to the Tropical Metabolism Research Unit (TMRU) ward at the University Hospital of the West Indies between the years 1963-1993 with a diagnosis of severe malnutrition. Members of the cohort were followed up and as adults and were extensively characterised in 2008-2012 in the Jamaica Marasmus and Kwashiorkor Adult Survivors (JAMAKAS) Study (**Figure 1**) [4, 13-15]. All adult severe malnutrition survivors followed up in the JAMAKAS Study for whom childhood hospital admission records were available were included in this analysis. The Mona Campus Research Ethics Committee of the University of the West Indies approved the study (CREC-MN. 204 20/21) and all participants provided written informed consent.

**Figure 1:**
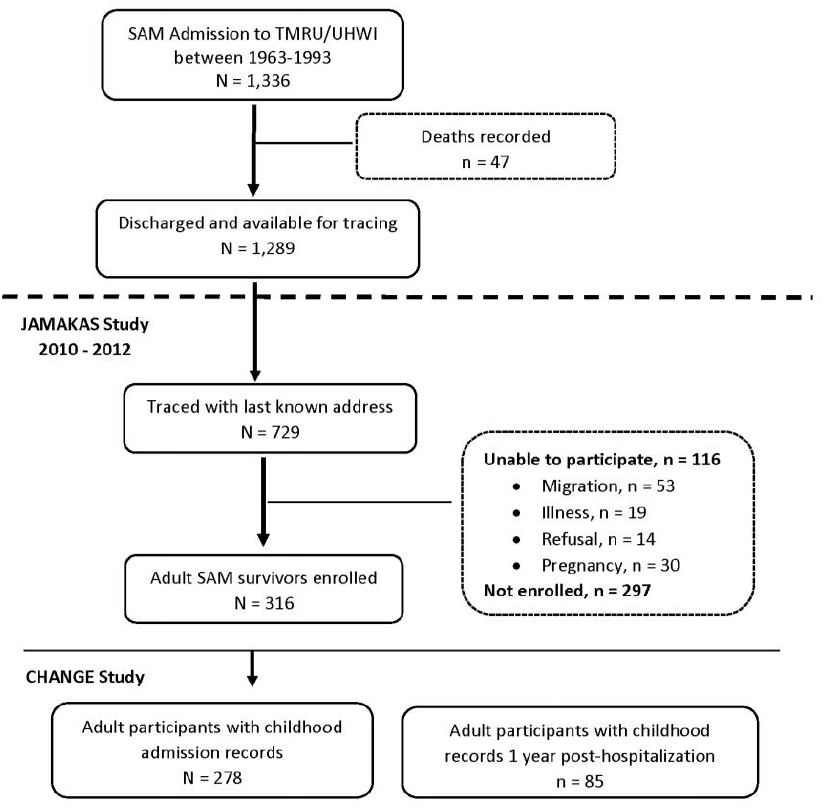
Flow chart detailing recruitment of adult survivors of severe malnutrition (*n* = 278). “Unable to participate” includes adult survivors of severe malnutrition who were unavailable because of migration (*n* = 53), illness (*n* = 19), refusal (*n* = 14), or pregnancy (*n* = 30). TMRU, Tropical Metabolism Research Unit; UHWI, University Hospital of the West Indies; JAMAKAS, Jamaica Marasmus and Kwashiorkor Adult Survivors.

During hospitalization for severe malnutrition, treatment began with a “stabilisation” phase during which a diet was offered to provide energy for weight maintenance while treating acute complications such as infections, mineral and fluid imbalances. Since the 1960s, metabolism research has informed development of treatment guidelines in the TMRU [16] which led to a gradual standardisation of the approach by about the mid-1970s [17]. Since then, the treatment regime has been similar to the guidelines by WHO [18]. With clinical improvement, resolution of oedema and improved appetite and affect, the “rehabilitation” phase occurred, during which children were fed increasing amounts of energy and protein-enriched feeds to attain a body weight similar to WHO (1981) guidelines [18]. The final phase of treatment, referred to as “recovery”, involved weaning the child to an age-appropriate diet before discharge from hospital. Children were discharged having attained weight-for-height ≥ 90% (NCHS) and were followed up as outpatients for up to two years post-hospitalization. During this period, they were fed a home-based diet.

Participant Characteristics: Childhood data abstracted from hospital records included admission weight, height and mid-upper arm circumference (MUAC) as well as weight, height/length and MUAC during hospitalization and one year post-discharge from hospital. Adult data were collected at the time of the JAMAKAS Study and included a detailed medical and drug history, anthropometry, blood pressure, body composition (by dual-energy x-ray absorptiometry, GE Lunar Prodigy), fasting glucose, insulin and serum lipids as described previously [19].

Exposure variables: We derived six definitions of post-malnutrition growth rate (PMGr) based on available data and established assessment methods (**Table 1**). The first three definitions (PMGr 1-3) were derived from in-patient weight and age data starting at the time of minimum weight, which was usually at admission or after oedema had resolved and continuing until the time of maximum weight during treatment. The definitions respectively utilized the daily change in weight-for-age z-score (WAZ) (using WHO 2006 growth standards) (PMGr1), g/kg of body weight (PMGr2) and weight in grams (PMGr3) (**Table 1**), all common measures of growth in malnutrition programs. Height-for-age z-score (HAZ) was not used during this period as height gain was small. Due to individual variations in the attainment of maximum weight, weight gain was expressed ‘per day’. The remaining three definitions (PMGr 4-6) represented weight gain and height gain from the time of hospital discharge until one year post-discharge. For these definitions, WAZ (PMGr4), g/kg (PMGr 5) and HAZ (PMGr 6) were expressed as rate of change per month (**Table 1**).

**Table 1:** Definitions of post malnutrition growth (PMGr)

| PMGr class | Abbreviated title | Definition | Time |
| --- | --- | --- | --- |
| 1 | $\Delta$ WAZ/day | Average change in WAZ per day | Date of minimum weight to date of maximum weight during hospitalization |
| 2 | g/kg/day | Average change in grams per kilogram of body weight per day |  |
| 3 | g/day | Average change in grams per day |  |
| 4 | $\Delta$ WAZ/month | Average change in WAZ per month | Date from hospital discharge to one year post-hospitalization |
| 5 | g/kg/month | Average change in grams per kilogram of body weight per month |  |
| 6 | $\Delta$ HAZ/month | Average change in HAZ per month | |

WAZ- weight-for-age z-scores, HAZ-height-for-age z-scores

Outcome variables included BMI, waist circumference, fat mass, fat mass index, % fat mass, android fat, android: gynoid (AG) fat ratio (android fat divided by gynoid fat; a pattern of body fat distribution associated with an increased risk for metabolic syndrome in healthy adults), lean mass, lean mass index, systolic blood pressure, diastolic blood pressure, fasting glucose, fasting insulin, LDL-cholesterol, triglyceride, and homeostatic model assessment for insulin resistance (HOMA-IR). Body composition (height-adjusted lean mass and fat mass) was assessed by dual-energy X-ray absorptiometry (DEXA).

Lean mass index based on DEXA was calculated using the formula: lean mass/height^2^, and similarly fat mass index based on DEXA was calculated using the formula: fat mass/height^2^, each in BMI units of kg/m^2^. HOMA-IR was calculated using the formula:

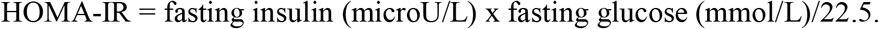

Statistical Analyses: To determine the relationship between PMGr and adult body size, body composition and NCD risk factors, we conducted separate linear regression analyses for each outcome adjusted for age and sex and (as an additional model) minimum WAZ (i.e., WAZ at the time of minimum weight). Additionally, all outcome measures of body composition and blood pressure were adjusted for adult height. Adult BMI was treated as an outcome variable and not adjusted for in the analyses as it was considered to be on the causal pathway between post malnutrition weight gain and the outcomes. Each PMGr definition was evaluated as a continuous variable, and we reported differences in continuous outcomes per unit faster growth depending on the definition of PMGr. Based on the six definitions of post malnutrition growth, participants were also grouped into quintiles which were used in age- and sex-adjusted regression analyses as ordinal variables. Quintiles 1 (Q1) and 5 (Q5) represented the slowest and fastest rates of growth respectively, and age- and sex-adjusted regression analyses were conducted comparing NCD risk across the growth rate quintiles. Finally, a latent class analysis (LCA) was conducted to identify qualitatively different growth pattern subgroups of the study population across 5 time points, viz., admission, time at minimum weight, time at maximum weight, discharge from hospital and one year post-hospitalization. The LCA was fitted using a generalised structural equation model with Gaussian-distributed outcomes at each of the timepoints included in the analysis. Model fit was assessed using Akaike Information Criteria (AIC); latent classes were extracted from the model with the lowest AIC, and which yielded reasonably-sized classes. These LCA-predicted classes were plotted against anthropometric, body composition and clinical markers of NCD risk to facilitate interpretation. Stata version 16.0 (StataCorp LLC, College Station, Texas, USA) was used to conduct the statistical analyses and *p*-values ≤ 0.05 were considered statistically significant.

## Results

There were 1,366 children in the original TMRU/UHWI cohort, 47 of whom died during treatment for malnutrition and the remaining 1,289 were discharged alive. The JAMAKAS study traced 729 adult survivors from the original cohort and enrolled 316 of them to the study. Subsequently, this current ‘CHANGE’ (Child malnutrition & adult NCD: Generating evidence on mechanistic links to inform future policy/practice) Study identified 278 adult participants for whom childhood admission records were available for inclusion to this study. **Figure 1** summarises the recruitment of the adult survivors of severe malnutrition.

We analysed data for 278 adults (60% male) who were treated for severe malnutrition as children. Mean age at admission was 10.9 months, 237/270 (88%) were stunted (HAZ < -2), 207/270 (77%) and 185/214 (87%) were wasted (WHZ <-2 and MUAC <12.5 cm respectively) and 182/278 (65%) had oedematous malnutrition. As adults, mean age (SD) was 28.2 (7.7) years, and mean BMI (SD) was 23.6 (5.2) kg/m^2^ (**Table 2**).

**Table 2:** Clinical characteristics of 278 survivors of severe malnutrition as children and as adults.

| Children | Characteristics | At Admission<br>N = 278 | At Discharge<br>n= 276 | At 1 year PH<br>n = 85 |
| --- | --- | --- | --- | --- |
|  | Age (months) | 11.9 (0.33 -36) | 13.7 (3 - 42) | 26.1 (15- 45) |
|  | % Male | (167) 60.1 % | 60.1% | 56.5% |
|  | % Oedema | (182) 65.5% | (0) 0% | (0) 0% |
|  | Weight (kg) | 5.4 (1.5), 278 | 7.3 (1.8), 276 | 9.8 (1.6), 85 |
|  | Height (cm) | 64.6 (7.0), 270 | 66.5 (7.2), 195 | 80.1 (5.6), 81 |
|  | MUAC (cm) | 10.8 (1.6), 214 | 12.9 (1.2), 33 | 14.(1.5), 18 |
|  | % Underweight (WAZ<-2) | 92 | 63 | 41 |
|  | % Stunted (HAZ<-2) | 88 | 88 | 59 |
|  | % Wasted (MUAC < 12.5cm) | 87 | 27 | 17 |
|  | % Wasted (WHZ < -2) | 77 | 5 | 15 |
|  | Days since admission | 0 | 55 (13 - 251) | 412 (343 - 588) |
| Adults | Characteristics | All<br>N = 278 | Male<br>n = 167 | Female<br>n= 111 |
|  | Years since discharge | 27.2 (16-51) | 27.3 (16-51) | 26.5 (17-46) |
|  | Age (years) | 28.2 (7.7) | 28.5 (7.9) | 27.7 (7.4) |
|  | Weight (kg) | 66.1 (14.9) | 65.5 (11.7) | 67 (18.7) |
|  | Height (cm) | 167.5 (9.0) | 171.1 (7.6) | 162 (7.8) |
|  | BMI (kg/m<sup>2</sup>) | 23.6 (5.2) | 22.3 (3.4) | 25.4 (6.7) |
|  | Waist circumference (cm) | 78.8 (12.7) | 76.3 (8.9) | 82.6 (16.2) |
|  | Waist-hip ratio | 0.83 (0.06) | 0.83 (.06) | 0.82 (0.07) |
|  | Diastolic blood pressure (mmHg) | 71 (11) | 71 (11) | 71 (12) |
|  | Systolic blood pressure (mmHg) | 114 (12) | 116 (12) | 112 (13) |
|  | Lean mass (kg) | 48.4 (10.4) | 54.4 (7.5) | 39.3 (6.9) |
|  | Lean mass index (kg/m<sup>2</sup>) | 17.1 (2.6) | 18.5 (2.0) | 14.9 (1.8) |
|  | Fat mass (kg) | 14.4 (13.2) | 8.0 (7.3) | 24.2 (14.1) |
|  | Fat mass index (kg/m<sup>2</sup>) | 5.3 (5.0) | 2.7 (2.5) | 9.2 (5.3) |
|  | % Fat | 21.2 (15.5) | 11.8 (8.6) | 35.4 (12.5) |
|  | Android fat mass (kg) | 1.1 (1.2) | 0.64 (0.68) | 1.9 (1.4) |
|  | Android fat % | 24.1 (17.5) | 14.4 (11) | 38.8 (15) |
|  | Android-gynoid fat ratio | 0.9 (0.22) | 0.9 (0.20) | 0.9 (0.24) |
|  | Fasting blood glucose (mmol/L)<sup>‡</sup> | 4.6 (0.5) | 4.7 (0.4) | 4.5 (0.5) |
|  | Fasting serum insulin (uIU/mL)<sup>‡†</sup> | 4.7 (3.6) | 3.3 (3.2) | 6.3 (3.4) |
|  | HOMA-IR<sup>†</sup> | 0.9 (0.7) | 0.7 (0.7) | 1.3 (0.7) |
|  | LDL-cholesterol<sup>†</sup> (mmol/L) | 2.1 (0.7) | 2.0 (0.6) | 2.3 (0.7) |
|  | Triglyceride<sup>†</sup> (mmol/L) | 0.7 (0.4) | 0.7 (0.3) | 0.7 (0.4) |

N = 278. PH = post-hospitalization. Z-scores based on WHO 2006 and 2007 references. Data are presented as means ( ± SDs) or (range). (<sup>‡</sup>, <sup>††</sup>, <sup>†</sup>) n = 95, 83 and 82 respectively.

Subsequent analyses were based on 273 participants during rehabilitation and 84 participants post-hospitalization as some participants had missing maximum weight or discharge weight data. In 273 participants, mean weight gain during rehabilitation was, according to the three definitions, 0.07 WAZ/day, 10.1 g/kg/day and 62 g/day (**Table 3**). Mean (SD) change in WAZ during rehabilitation was 2.2 (0.97). In the post-hospitalization phase, weight gain was much lower; mean (SD) weight gain was 0.8 (0.5) g/kg/day and mean (SD) change in WAZ/day was 0.002 (0.004) for 84 participants. Each PMGr definition was grouped in quintiles (**Table 3 and Figure 2**)

**Table 3:** Descriptive data for definitions of post-malnutrition growth (PMGr)

| PMGr Class | Description | Mean (SD) | Range |
| --- | --- | --- | --- |
| **1**<br>(n = 273) | **Δ WAZ/ day** | 0.07 (0.03) | -0.0025, 0.20 |
|  | Q1 | 0.03 (0.01) | -0.00, 0.04 |
|  | Q2 | 0.05 (0.00) | 0.04, 0.06 |
|  | Q3 | 0.06 (0.00) | 0.06, 0.07 |
|  | Q4 | 0.08 (0.00) | 0.07, 0.09 |
|  | Q5 | 0.11 (0.02) | 0.09, 0.20 |
| **2**<br>(n = 273) | **g/kg/day** | 10.1 (3.8) | 0.32 - 27 |
|  | Q1 | 5.5 (1.3) | 0.3, 7.0 |
|  | Q2 | 7.9 (0.6) | 7.0, 8.9 |
|  | Q3 | 9.7 (0.5) | 8.9, 10.7 |
|  | Q4 | 11.8 (0.6) | 10.7, 12.9 |
|  | Q5 | 15.7 (2.9) | 12.9, 27.4 |
| **3**<br>(n = 273) | **g/day** | 61.6 (25.3) | 2.5 - 189 |
|  | Q1 | 31.8 (8.0) | 2.5, 41.0 |
|  | Q2 | 46.9 (3.1) | 41.4, 51.7 |
|  | Q3 | 58.6 (3.9) | 52.0, 64.8 |
|  | Q4 | 71.6 (5.1) | 65.0, 81.0 |
|  | Q5 | 99.9 (20.5) | 81.1, 188.7 |
| **4**<br>(n = 84) | **Δ WAZ/ month** | 0.05 (0.1) | -0.22 - 0.46 |
|  | Q1 | -0.09 (0.06) | -0.22, -0.03 |
|  | Q2 | -0.01 (0.01) | -0.03, 0.01 |
|  | Q3 | 0.05 (0.02) | 0.01, 0.08 |
|  | Q4 | 0.11 (0.02) | 0.08, 0.15 |
|  | Q5 | 0.21 (0.08) | 0.15, 0.46 |
| **5**<br>(n = 84) | **g/kg/month** | 24.9 (15.6), | -0.5 - 7.1 |
|  | Q1 | 5.4 (8.0) | -12.9, 14.4 |
|  | Q2 | 16.9 (2.4) | 14.6, 20.9 |
|  | Q3 | 24.5 (2.2) | 21.4, 28.0 |
|  | Q4 | 32.0 (3.3) | 28.1, 37.2 |
|  | Q5 | 47.2 (12.9) | 37.8, 90.2 |
| **6**<br>(n = 47) | **Δ HAZ/ month** | 0.10 (0.12) | -0.08 - 0.54 |
|  | Q1 | -0.04 (0.03) | -0.08, 0.01 |
|  | Q2 | 0.04 (0.01) | 0.02, 0.06 |
|  | Q3 | 0.09 (0.01) | 0.07, 0.11 |
|  | Q4 | 0.14 (0.02) | 0.12, 0.17 |
|  | Q5 | 0.29 (0.11) | 0.18, 0.54 |

PMGr - post malnutrition growth rate, Q – quintile: 1- slowest growth to 5- fastest growth, WAZ - weight-for-age z-scores, HAZ - height-for-age z-scores

**Figure 2:**
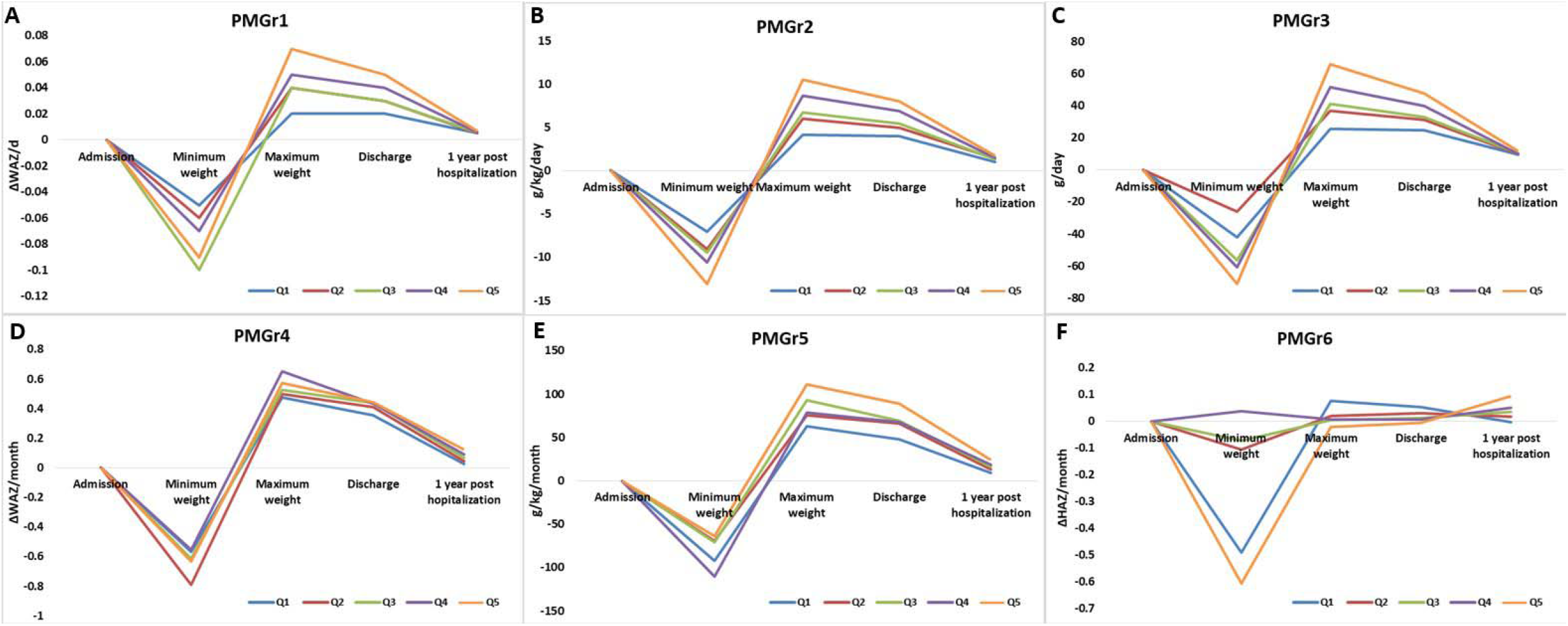
Mean growth rates of growth across four time periods according to the six PMGr definitions. A to C: PMGr1 to 3 (*rehabilitation*) and D to F: PMGr4 to 6 (*post-hospitalization*). Each PMGr definition was split into quintiles where Q1= slowest growth, and Q5 = fastest growth. PMGr at admission acted as comparator for the later timepoints.

### Associations between rehabilitation weight gain and adult NCD risk

The three measures of rehabilitation weight gain (n = 273) were associated with adult NCD risk in regression models adjusted for age, sex and minimum WAZ as follows:

**PMGr1** as ΔWAZ/day was associated with waist circumference (difference=53cm, 95% CI: 6, 100, *p* = 0.03) and lean mass (difference=37.0 kg, 95% CI: 7, 67, *p* = 0.02) (**Supplementary Table 1**). Those with fastest weight gain had higher BMI (difference = 0.4kg/m^2^, 95% CI: 0.02, 0.8, *p* = 0.04), waist circumference (difference = 1.2 cm, 95% CI: 0.3, 2, *p* = 0.01), lean mass (difference = 0.8 kg, 95% CI: 0.2, 1.4, *p* = 0.01), fat mass (difference = 0.8 kg, 95% CI: 0.01, 1.7, *p* = 0.048) and % fat mass (difference = 0.85, 95% CI: 0.02, 1.7, *p* = 0.04) than those with slowest weight gain (**Supplementary Table 2, Figure 3)**. Associations with waist circumference and % fat mass remained significant after further adjusting for adult height. Visual examination of the box plots **(Figure 3A, 3C, 3K, 3N)** suggested that only ΔWAZ ≥ 0.09/day was associated with measures of adult adiposity and when further exploratory analyses (excluding Q5) were conducted, ΔWAZ < 0.09/day was not associated with adult adiposity (*p-*values > 0.2) (data not shown). In the fastest weight gainers (Q5) in comparison to the slowest weight gainers (Q1), ΔWAZ/day was associated with BMI (difference = 2.1 kg/m2, 95% CI: 0.3, 3.8, *p* = 0.02), waist circumference (difference = 6.5 cm, 95% CI: 2, 11, *p* = 0.002), waist-hip ratio (difference = 0.03, 95% CI: 0.003, 0.05, *p* = 0.03), fat mass (difference = 4.0 kg, 95% CI: 0.2, 8, *p* = 0.04) and android fat mass (difference = 0.36 kg, 95% CI: 0.01, 0.7, *p* = 0.05) (data not shown).

**Figure 3:**
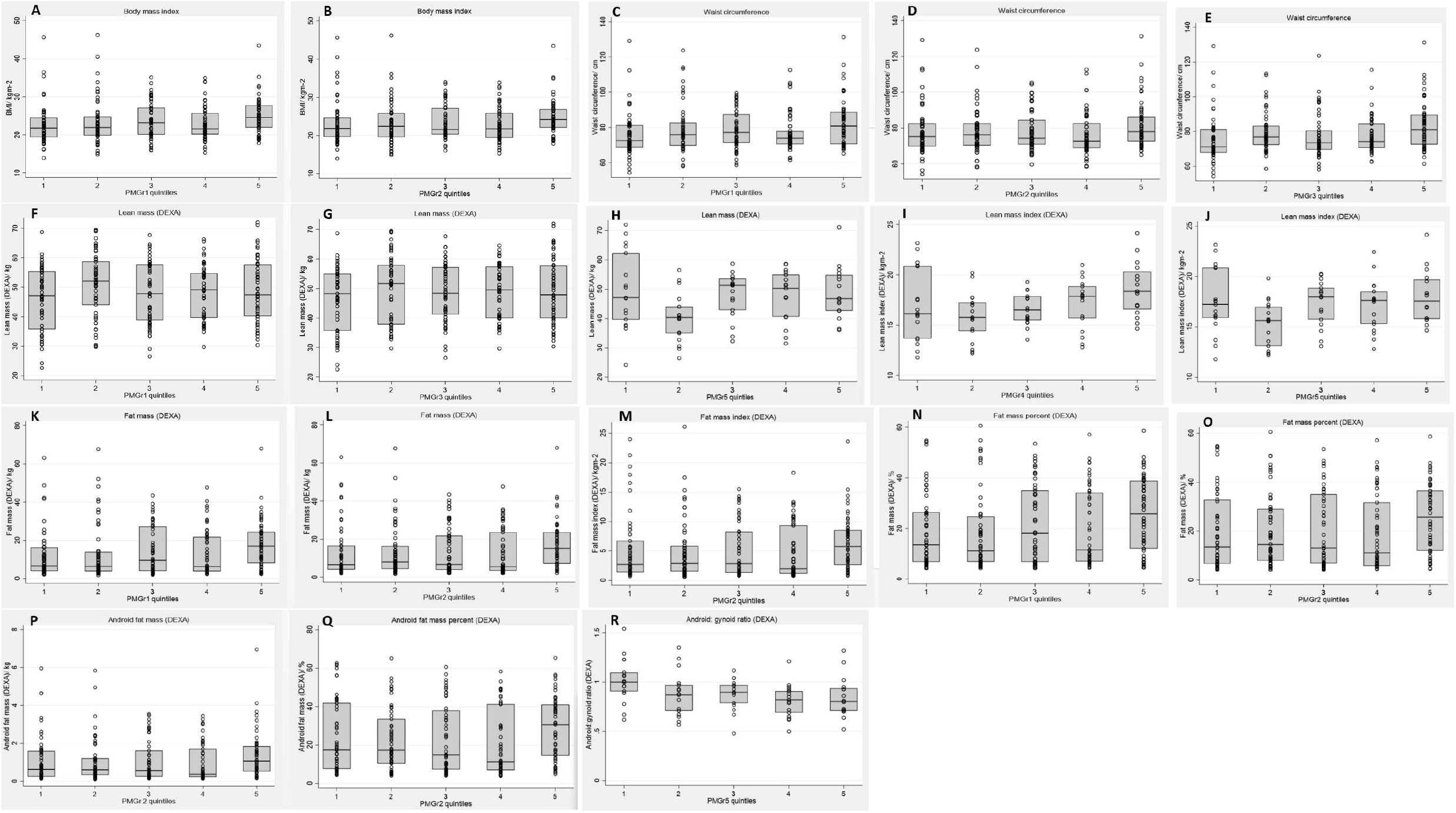
Association of NCD outcomes with PMGr definitions. Age, sex and minimum weight WAZ-adjusted linear regression results (coefficient, p-value): **A to B**-body mass index: PMGr1 (B = 0.4, p = 0.04), PMGr2 (B = 0.5, p = 0.02); **C to E**-waist circumference: PMGr1 (B = 1.2, p = 0.01), PMGr2 (B = 1.4, p = 0.01), and PMGr3 (B = 1.2, p = 0.01); **F to H**-lean mass: PMGr1 (B = 0.8, p = 0.01), PMGr3 (B = 0.7, p = 0.02) and PMGr5 (B = 1.3, p = 0.02); **I to J**-lean mass index: PMGr4 (B = 0.35, p = 0.03) and PMGr5 (B = 0.43, p = 0.01); **K to L**-fat mass: PMGr1 (B = 0.84, p = 0.048) and PMGr2 (B = 1.1, p = 0.01); **M**-fat mass index: PMGr2 (B = 0.32, p = 0.05); **N to O**-fat mass percent: PMGr1 (B = 0.85, p = 0.04) and PMGr2 (B = 1.1, p = 0.02); P-android fat mass: PMGr2 (B = 0.09, p = 0.03); **Q**-android fat mass %: PMGr2 (B = 1.2, p = 0.03); **R**- android-gynoid ratio: PMGr5 (B = -0.03, p = 0.045).

**PMGr2** as weight gain in g/kg/day was associated with waist circumference (difference = 0.43 cm, 95% CI: 0.1, 0.8, *p* = 0.02) and lean mass (difference = 0.28 kg, 95% CI: 0.05, 0.5, *p* = 0.02) (**Supplementary Table 1**). Those with fastest weight gain had higher BMI (difference = 0.5 kg/m^2^, 95% CI: 0.1, 0.9, *p* = 0.02), waist circumference (difference = 1.4 cm, 95% CI: 0.4, 2.4, *p* = 0.005), fat mass (difference = 1.1 kg, 95% CI: 0.2, 2, *p* = 0.02), fat mass index (difference = 0.32 kg/m2, 95% CI: -0.0001, 0.6, *p* = 0.05), % fat mass (difference = 1.1, 95% CI: 0.2, 2, *p* = 0.02), android fat mass (difference = 0.09 kg, 95% CI: 0.01, 0.2, *p* = 0.03) and % android fat mass (difference = 1.2, 95% CI: 0.1, 2, *p* = 0.03) than those with slowest weight gain (**Supplementary Table 2, Figure 3**). The associations with waist circumference, % fat mass, android fat mass and % android fat mass remained significant after further adjusting for height (data not shown). Examination of the box plots **(Figure 3B, 3D, 3L, 3M, 3O, 3P, 3Q)** suggested that only weight gain ≥ 12.9 g/kg/day was associated with measures of adult adiposity and when further exploratory analyses (excluding Q5) were conducted, rehabilitation weight gain <12.9 g/kg/day was not associated with adult adiposity (*p-*values > 0.3) (data not shown).

**PMGr3** as weight gain in g/day was associated with lean mass (difference = 0.04 kg, 95% CI: 0.01, 0.1, *p* = 0.02) (**Supplementary Table 1**). Those with fastest weight gain had greater waist circumference (difference = 1.2 cm, 95% CI: 0.2, 2, *p* = 0.01) and lean mass (difference = 0.7 kg, 95% CI: 0.1, 1.3, *p* = 0.02) than those with slowest weight gain (**Supplementary Table 2, Figure 3**). Examination of the box plots **(Figure 3E)** suggested that rehabilitation weight gain ≥ 81 g/day was associated with adult waist circumference and when further exploratory analyses were conducted, weight gain < 81 g/day was not associated with adult waist circumference (*p-*values > 0.3) (data not shown).

There was no association between any measure of rehabilitation weight gain (i.e., PMGr1, PMGr2, and PMGr3) and adult blood pressure, blood glucose, insulin, or lipids **(Supplementary Tables 1 and 2)**.

### Associations between post-hospitalization weight gain and height gain and adult NCD risk

Post-hospitalization weight gain (n = 84) and height gain (n = 47) were associated with NCD risk in regression models adjusted for age, sex and minimum WAZ as follows:

**PMGr4** as WAZ/month was associated with lean mass index (difference = 4.6 kg/m^2^, 95% CI: 0.6, 9, *p* = 0.03) (**Supplementary Table 1**). Those with fastest weight gain had a higher lean mass index (difference = 0.4 kg/m^2^, 95% CI: 0.04, 0.7, *p* = 0.03) than those with the slowest weight gain (**Supplementary Table 2, Figure 3**).

**PMGr5** as g/kg/month was associated with lean mass index (difference = 0.03 kg/m^2^, 95% CI: 0.01, 0.06, *p* = 0.02) (**Supplementary Table 1**). The fastest weight gainers had higher lean mass (difference = 1.3 kg, 95% CI: 0.3, 2.4, *p* = 0.02) and lean mass index (difference = 0.4 kg/m^2^, 95% CI: 0.1, 0.7, *p* = 0.03) and a lower AG ratio (difference = -0.03, 95% CI: -0.07, -0.001, *p* = 0.05) than the slowest weight gainers (**Supplementary Table 2, Figure 3**).

**PMGr6** as height gain in ΔHAZ/month was not associated with NCD outcomes and there was no difference between the fastest and slowest growers in terms of the NCD outcomes.

There was no association between any measure of post-hospitalization weight gain or growth (i.e., PMGr4, PMGr5, and PMGr6) and adult blood pressure, blood glucose, insulin, or lipids **(Supplementary Tables 1 and 2)**.

### Latent class analysis

Latent class analysis (LCA) identified a model with 6 interpretable classes of at least 21 children each which had the lowest AIC (3159) (**Figure 4**). Class 1 (10%) consisted of the children who were most underweight on admission (mean WAZ < -6) and with the least weight gain during rehabilitation. Class 2 (21%) had the second most underweight children with the second lowest rehabilitation weight gain. Class 3 (30%) were children with the second highest rehabilitation weight gain. Class 4 (18%) had the highest rehabilitation weight gain. Class 5 (13%) had the second highest WAZ at admission and the third highest rehabilitation weight gain. Class 6 (8%) were the children who were least underweight on admission. All classes have a similarly steep level of catch-up during rehabilitation, except class 4. After discharge, class 1, with the greatest deficits, continued to catch up, and there was a pattern of decreasing catch-up growth up the classes, until class 6 where “catch down” was observed (**Figure 4)**.

**Figure 4:**
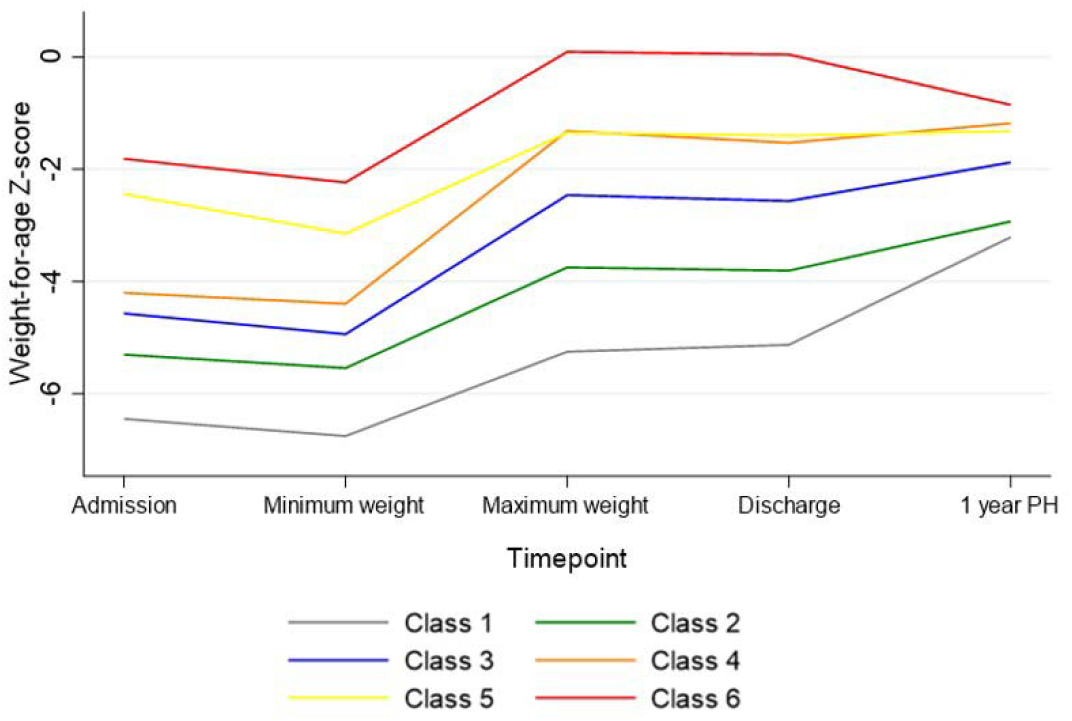
Latent class analysis of patterns of weight-for-age z-score from admission for severe malnutrition to 1 year post-hospitalization in 278 adults. Classes ranked by lowest WAZ upon admission: 1, 2, 3, 4, 5, 6, and by highest weight gain during rehabilitation: 4, 3, 6, 5, 2, 1. PH-post hospitalization

Age and sex-adjusted regression analyses against the NCD outcomes were carried out with LCA class as a categorical variable and Class I being the reference group. In children in Class 5 (of which 89% had oedematous malnutrition), change in WAZ was associated with greater adult fat mass (difference = 5.2kg, SE = 2.6; 95% CI: 0.1, 10, *p* = 0.045) and android fat mass (difference = 0.52 kg, SE = 0.24; 95% CI: 0.04, 0.99, *p* = 0.03) compared to children in Class 1. However, these associations were no longer significant after adjusting for minimum WAZ (data not shown).

## Discussion

This study of adult survivors of severe malnutrition in childhood demonstrated that, during the rehabilitation phase of malnutrition treatment, weight gain above certain thresholds is associated with risk factors for NCDs (i.e., greater BMI, waist circumference, fat mass and android fat mass). Weight gain during the first year after hospitalization was inversely associated with android-gynoid fat ratio. Additionally, in adult survivors, faster weight gain during and after admission for severe malnutrition was associated with lean mass which is typically associated with lower NCD risk. It is important to note that these associations are independent of weight-for-age at the time of hospitalization and adult height. Neither rehabilitation weight gain nor post-hospitalization weight gain was associated with adult blood pressure or blood glucose, insulin, or lipids.

### Post-malnutrition growth and adult adiposity

Weight gain targets during treatment of severe malnutrition have been in place since the 1970s, with the recommendation that a desirable rehabilitation weight gain exceeds 10g/kg/day [20]. We have demonstrated in this group of adult survivors of severe malnutrition that rehabilitation weight gain that exceeded specified thresholds was associated with adult adiposity. Measures of adiposity (i.e., BMI, waist circumference, fat mass and android fat mass) appeared similar across the first four quintiles of weight gain (defined as ΔWAZ, g/kg/day and g/day), whereas the fifth quintile showed greater adiposity. Thus, each unit increase in weight gain does not appear to increase NCD risk in the same way, and there is a threshold effect of rehabilitation weight gain on adult adiposity. This suggests that weight gain at or above specific thresholds (i.e., 13 g/kg/day, ΔWAZ > 0.09/day, 81g/day) could impute greater adult adiposity.

Faster rehabilitation weight gain was associated with greater adult BMI (an indicator of excess or inadequate weight) which itself is an independent risk factor for cardiovascular disease. Weight gain above the previously specified thresholds was also associated with increased fat mass, and this excess adipose tissue contributes to the production of proinflammatory cytokines which are also associated with increased risk of cardiovascular disease. Importantly, faster rehabilitation weight gain was associated with fat mass index, which has been shown to be a more complete tool for diagnosing obesity than both BMI and fat mass as it differentiates fat and muscle mass of the total body weight [21]. Furthermore, fat accumulation in the android compartment is specifically known to be associated with increased cardiovascular and metabolic risk [22] and studies in younger populations have also demonstrated that android fat is more closely related to metabolic risk factors [23]. This tendency for some adult survivors of severe malnutrition to accumulate visceral fat may be a consequence of adipose tissue maldevelopment which includes altered body composition and increased visceral fat [24]. Rapid postnatal infant weight gain has been shown to be associated with increases in both visceral and abdominal subcutaneous adipose tissue, as well as total adiposity and the risk of obesity in middle adulthood [25]. Additionally, increased visceral adipose tissue is linked to high triglycerides and low HDL cholesterol levels [26]. However, while the tendency to store fat abdominally might be a persisting response to adverse conditions and growth failure in fetal life and infancy, we have shown this tendency to store fat in the abdominal compartment to be independent of minimum weight-for age z-score during hospitalization. Therefore, the association between post-malnutrition weight gain in childhood and adult android fat in this group of young, normal-weight severe malnutrition survivors could signal an increased risk of clinical NCDs as they age.

It is also important to point out that although our findings indicate statistically significant associations across the range of weight and body composition markers of NCD risk, these individuals have not exceeded established thresholds for adverse outcomes such as obesity. So, while faster weight gain was associated with higher adult BMI, only 11% of these participants were obese (i.e., BMI >30 kg/m^2^). Additionally, several studies have reported a lower susceptibility to visceral adipose tissue deposition in blacks compared to whites [27, 28]. Therefore, although it is unclear whether the levels of visceral fat in these study participants are sufficient to impute cardiometabolic risk, the possibility cannot be ruled out.

Our study produced the unexpected finding of an inverse association between post-hospitalization weight gain (g/kg/month) and android-gynoid percent fat ratio (AG), an important predictor of metabolic and cardiovascular disease risk. Indeed, in over 1,800 participants with a mean age of 35 years, AG was reported to have a greater association with cardiometabolic dysregulation than android fat, gynoid percent fat or BMI [29]. While our findings are counterintuitive, they further emphasize the complexity of these associations which may depend not only on the extent and rate of weight gain, but also the age of the child at the time of weight gain, and other factors outside the scope of this study.

### Post-malnutrition growth and adult lean mass

Lean mass in adult survivors of severe malnutrition was associated with both rehabilitation and post-hospitalization weight gain. While greater lean mass is generally associated with reduced NCD risk, the association of lean mass with health appears complex [30]. Although lean mass incorporates muscle mass, which is widely considered to protect against diabetes, high levels of lean mass have been associated with higher blood pressure [31]. Additionally, it is possible that in this group, lean mass is accreted in tandem with fat mass, with the benefit of higher lean mass being obscured by the deleterious effects of fat (particularly visceral fat) accumulation.

### The role of minimum WAZ

It is important to establish the role of minimum weight-for-age in these analyses as it has been reported that the most underweight children tend to gain more weight during nutritional rehabilitation. In our participants, however, most of the reported associations between both rehabilitation and post-hospitalization weight gain and adult adiposity were independent of minimum WAZ. However, in one subgroup of participants identified by latent class analysis, minimum WAZ appears to be influential. Therefore, the relative importance of rehabilitation weight gain versus admission WAZ in predicting NCD risk in these adult survivors may be dependent on how weight gain is defined, and there could be an interplay between these two factors influencing the association between childhood weight gain and adult adiposity.

### Post malnutrition growth rate

Rapid weight gain in infancy has been defined as a weight-for-age Z score gain greater than 0.67 between birth and 1.5 years [32]. Others have defined it as a change in WAZ > 0.67 over an unspecified time period [33, 34]. Our study participants had a mean change in WAZ during rehabilitation (mean duration 37 days) that was more than 3 times the + 0.67 cut-off. Our statistical observation of associations between a one-unit change in WAZ per day (ΔWAZ/day) and very large increases in lean mass and waist circumference must be interpreted with the caution that such an increase in WAZ is biologically implausible. Nevertheless, our findings call into question current guidelines which also recommend that a weight gain of >10 g/kg/day during rehabilitation is desirable, as some of our participants who gained more than 10/g/kg/day during rehabilitation were shown to have greater adiposity as adults. In India, WAZ and g/kg/day disagreed substantially as methods for measuring growth velocity in more than a third of preterm children studied [35]. How the two measures agree in children born at term is unclear, however, among our participants the two measures were similarly associated with measures of adult adiposity but not lean mass.

In summary, all three measures of in-treatment rehabilitation weight gain were associated with at least one measure of adult adiposity; the same was not true of the measures of post-hospitalization weight gain. While z-score is considered the most appropriate descriptor of malnutrition, among our participants rehabilitation weight gain in g/kg/day had the most consistent associations with adult NCD risk factors and has the additional advantage of being more applicable in clinical settings. Nevertheless, we were unable to draw conclusions regarding the most appropriate proxy for post-malnutrition weight gain from this study.

## Strengths and Limitations

The strengths of the study include the uniqueness of the cohort and the availability of detailed anthropometric and body composition measures in children and adults. The low mortality (approximately 4%) minimizes the risk of survivor bias which limits some other malnutrition-survivor studies [36]. We acknowledge however that many post-hospitalization factors occurring in childhood, including the home diet and intercurrent illnesses, could have influenced the observed associations. Additionally, data relating to adult dietary intake, physical activity, and co-morbid conditions, all of which may confound the observed associations, were not evaluated. Finally, the participants in this study were Afro-Caribbean and the findings may be different in other ethnic groups and in other settings where other environmental and dietary conditions apply.

## Conclusion

In this unique cohort of adult Afro-Caribbean severe malnutrition survivors, we demonstrated statistically significant associations between rehabilitation and post-hospitalization weight gain and adult NCD risk that were independent of admission weight and adult height. While the implications of some of the findings are mixed, we have presented evidence of a risk of increased BMI and adiposity, especially abdominal adiposity, in these adults whose rehabilitation weight gain exceeded certain thresholds. Our findings raise the need to further explore the existing guidelines relating to weight gain targets during malnutrition treatment in childhood and underscore the need for further characterisation of optimal post-malnutrition weight gain in an interventional trial.

## Data Availability

All data produced in the present study are available upon reasonable request to the authors

## Acknowledgements

We gratefully acknowledge the men and women who took part in the study. We also recognize support from the members of the CHANGE Study Collaborators’ Group.

## Financial Support Statement

This work was supported by the UK Medical Research Council (grant number MR/V000802/1). The initial project was supported by the New Zealand Health Research Council Grant 09/052, Developmental Adaptation to an Obesogenic Environment Program.

## Notes

### Competing Interest Statement

The authors have declared no competing interest.

### Funding Statement

This study was funded by the UK Medical Research Council (grant number MR/V000802/1). The initial project was supported by the New Zealand Health Research Council Grant 09/052, Developmental Adaptation to an Obesogenic Environment Program.

### Author Declarations

The Mona Campus Research Ethics Committee of the University of the West Indies gave ethical approval for this work.

### Summary of Updates

The recommended energy intake during the three day transition period is 100-135 kcal/kg/day [7] followed by intake of 150-225 kcal/kg/d during the rehabilitation (catch-up growth) period, the goal being a weight gain of ≥ 10 g/kg/day as recommended by the World Health Organization (WHO) [8].

